# Different Associations of Anthropometric Indices with Diabetic Retinopathy and Diabetic Kidney Disease in Chinese Patients with Type 2 Diabetes Mellitus

**DOI:** 10.1101/2022.10.07.22280304

**Authors:** Yujie Wang, Xin Pang, Chufeng Gu, Chenxin Li, Bo Li, Chuandi Zhou, Haibing Chen, Zhi Zheng

## Abstract

**Aims:** To investigate the associations of anthropometric indices, including body mass index (BMI), waist-to-hip ratio (WHR), waist-to-height ratio (WHtR), waist circumference (WC) and hip circumference (HC), with diabetic retinopathy (DR) and diabetic kidney disease (DKD) in Chinese patients with type 2 diabetes mellitus (T2DM).

**Materials and methods:** This cross-sectional study evaluated 5226 participants with T2DM at Shanghai General Hospital between 2005 and 2016. Logistic regression models and restricted cubic spline analysis were used to assess the associations of anthropometric indices with DR and DKD.

**Results:** A BMI of around 25 kg/m^2^ was related to a low risk of DR (OR based on the third fifth: 0.752, 95%CI: 0.615-0.920). Besides, HC had an inverse association with DR in men independently of BMI (OR based on the highest fifth: 0.495, 95%CI: 0.350-0.697). In the restricted cubic spline models, BMI, WHtR, WC, and HC showed J-shaped associations with DKD, while WHR showed an S-shaped association with DKD. Compared to the lowest fifth, the odds ratios (OR) based on the highest fifth of BMI, WHR, WHtR, WC and HC for DKD were 1.927 (1.572-2.366), 1.566 (1.277-1.923), 1.91 (1.554-2.351), 1.91 (1.554-2.351) and 1.585 (1.300-1.937) respectively in multivariable models.

**Conclusions:** A median BMI and a large hip might be related to a low risk of DR, while lower levels of all the anthropometric indices were associated with a lower risk of DKD. Our findings suggested maintain a median BMI, a low WHR, a low WHtR and a large hip for prevention of DR and DKD.

## 1. Introduction

The global prevalence of diabetes has increased over the past decades and is expected to rise from an estimated 422 million people in 2014 to 783 million by 2045 ^12^. Type 2 diabetes mellitus (T2DM) accounts for around 90% of all diabetes cases. Diabetic retinopathy (DR) and diabetic kidney disease (DKD) are the major microvascular complications of diabetes, which can greatly reduce the quality of life and place a significant burden on health care costs. DR is the leading cause of vision loss and blindness worldwide. More than 60% of patients will eventually develop DR after 20 years of T2DM ^3^. DKD can develop in about 40% of patients with diabetes, leading to an increased risk of frailty, end-stage renal disease (ESRD), and premature mortality ^4^. Therefore, early detection and treatment of microvascular complications of diabetes is an important public health problem.

DR and DKD are usually comorbid, and they share some common pathogenic mechanisms, such as angiogenesis, oxidative stress and inflammation ^5^. However, there are apparent differences between the pathophysiologic features of DR and DKD: DR is characterized by vascular permeability and retinal neovascularization, while DKD is featured with glomerular sclerosis. It has been revealed that the associations of several clinical indicators with DR and DKD are different, for example, C peptide ^6^, glycated albumin to glycated hemoglobin (HbA1c) ratio ^7^, and age ^8^.

Obesity has been established as a risk factor for T2DM ^9,10^. However, the associations of obesity-related anthropometric indices, including body mass index (BMI), waist-to-hip ratio (WHR), waist-to-height ratio (WHtR), waist circumference (WC) and hip circumference (HC), with DR and DKD have not been systemically reported. According to our knowledge, evidence for these anthropometric indices with DR and DKD has been inconclusive. For instance, conflicting results were reported for the associations of BMI, WC, WHR and WHtR with DR, as positive ^11,12,13^, inverse ^14,15,16^ and no associations ^17^. Although numerous studies suggested that BMI, WC, WHR and WHtR were associated with a greater likelihood of having DKD ^18^, one study found that WC, WHR and WHtR had no significant associations with DKD ^19^.

Heterogeneous results were observed in previous studies where obesity-related parameters were analyzed as continuous variables or based on different categories ^19^, which probably indicated the existence of a non-linear trend. However, only a few studies were conducted to specifically analyze the non-linear associations between anthropometric indices and DR and DKD.

In this study, we aim to assess the associations of obesity-related anthropometric indices, including BMI, WHR, WHtR, WC and HC, with the prevalence of DR and DKD in 5226 Chinese individuals with T2DM.

## 2. Materials and methods

### 2.1 Study design and participants

This cross-sectional study was designed to investigate the associations of BMI, WHR, WHtR, WC and HC with DR and DKD among Chinese adults. Participants in this study were patients diagnosed with T2DM at Shanghai General Hospital between 2005 and 2016. Patients who were missing weight, height, WC or HC measurement or missing triglycerides (TG), high-density lipoprotein (HDL), low-density lipoprotein (LDL), HbA1c or systolic blood pressure (SBP) data or missing DR assessment information, or missing urinary albumin: creatinine (UACR) or estimated glomerular filtration rate (eGFR) information were excluded. Overall, 5226 patients were involved in this analysis.

The study was performed following the guidelines of the World Medical Association Declaration of Helsinki and was approved by the Institutional Review Board of the Shanghai General Hospital, Shanghai Jiao Tong University School of Medicine. Written informed consent was obtained from all patients or their legal guardians.

### 2.2 Data collection

The information on sociodemographic characteristics and medical history was collected by trained doctors through a standardized interview.

Height and weight were measured in cm and kg respectively with participants standing without shoes and in lightweight clothes. WC (in cm) was measured on the mid-axillary line between the lowest border of the rib cage and the top of the iliac crest. HC (in cm) was measured at the widest part of the hip at the level of the greater trochanter. BMI was calculated as weight in kilograms divided by squared height in meters. WHR was calculated as waist circumference divided by hip circumference. WHtR was calculated as waist circumference divided by height.

SBP and diastolic blood pressure (DBP) values were measured with participants in a sitting position after 10 minutes of rest.

Venous blood samples were drawn between 6:00 am and 9:00 am after an overnight fast. HbA1c, TC, TG, LDL, HDL, fasting blood glucose (FBG) and serum creatinine were assessed using a conventional automated blood analyzer. The 24-hour urine samples were collected during the period of hospitalization. The concentration of urine albumin and creatinine were measured with a turbidimetric immunoassay and an enzymatic method in a single 24-hour urine sample, respectively. Then the UACR was calculated.

### 2.3 Definition of Variables

Hypertension was defined as SBP≥140 mmHg, DBP≥90 mmHg, or a self-reported previous diagnosis of hypertension.

The eGFR was calculated according to the Chronic Kidney Disease Epidemiology Collaboration (CKD-EPI) equation for “Asian origin”^20^. The definition of DKD was UACR ≥30 mg/g or eGFR<60 mL/min*1.73 m^2, as suggested by the statement from the American Diabetes Association^21^.

DR screening was conducted by experienced ophthalmologists, including a review of ophthalmologic history, measurement of visual acuity and intra-ocular pressure, slit lamp examination, and dilated fundus examination. The digital retinal photography was undertaken after pupil dilation by a digital retinal camera (Carl Zeiss Meditec AG, Jena, Germany), and the Optical coherence tomography was obtained by Spectralis OCT (Heidelberg Engineering, Heidelberg, Germany). Retinopathy was graded based on the worst eye by well-trained assessors according to the Early Treatment Diabetic Retinopathy Study grading system ^22^.

### 2.4 Statistical analysis

Data analyses were performed with R 4.1.2 version. Continuous variables were expressed as the mean ± standard deviation (SD) or the median with an interquartile range (25%, 75%), and categorical variables were presented as percentages (%). Baseline characteristics were compared using the Chi-square test for dichotomous variables, the Mann-Whitney U test or the Student’s t test for continuous variables as appropriate. P value (two-sided) <0.05 indicated significance.

We calculated a Spearman correlation between every two variables of BMI, WHR, WHtR, WC and HC (Table S1). We used binomial logistic regression models to analyze the associations of these anthropometric indices with DR and DKD. BMI, WHR, WHtR, WC and HC were analyzed continuously and categorically in fifths. Data were summarized as odds ratios (OR) and 95% confidence intervals (95%CI). For the primary analysis, we conducted three adjusted regression models: model 1 adjusted for age and sex; model 2 adjusted for age, sex, duration of diabetes, SBP, TG, LDL, HDL and HbA1c; model 3 further including BMI for analyses of WHR, WHtR, WC and HC or including WHR for analyses of BMI. Ordinal logistic regression models were used to assess the overall trend of categorical exposure with the presence of DR and DKD.

We also used restricted cubic splines to examine the shape of the associations of BMI, WHR, WHtR, WC and HC with DR and DKD. The numbers of knots were set between 3 and 5 based on Akaike’s information criteria ^23^. The locations of knots were prespecified based on the quantiles of the continuous variable (Table S2). The median values of these variables were used as references. Age, sex, duration of diabetes, SBP, TG, LDL, HDL and HbA1c were adjusted in the spline models. P for non-linearity was tested with the Wald test, and a P value<0.05 was considered to show a significant non-linear trend. We also used a linear model to calculate OR per SD increase in a specific range of these variables.

Because evidence indicated a significant interaction between sex and BMI, WHtR, WC or HC with DR and DKD (table S3), we conducted sex-stratified analyses of all these anthropometric indices with the presence of DR and DKD.

We tested the consistency of our findings by using different categories for BMI. And we also did several sensitivity analyses of WC and HC with DR in men with additional adjustments for WHR, WC or HC.

## 3. Results

### 3.1 General characteristics of study participants by DR and DKD

We included 5226 patients with T2DM in this analysis. Overall, the mean age was 59.2 ± 12.0 years and of these patients, 1662 (31.8%) had DR and 1742 (33.3%) had DKD.

Table1 summarizes the clinical and demographic characteristics of the study population in those with or without DR, and with or without DKD. No difference in BMI, WHtR and WC were found between patients with and without DR (all P >0.05). And patients with DR were likely to have a larger WHR and a smaller HC than those without DR (P <0.05). However, compared with patients without DKD, BMI, WHR, WHtR, WC and HC were significantly higher in those with DKD (all P <0.01).

**Table 1.**
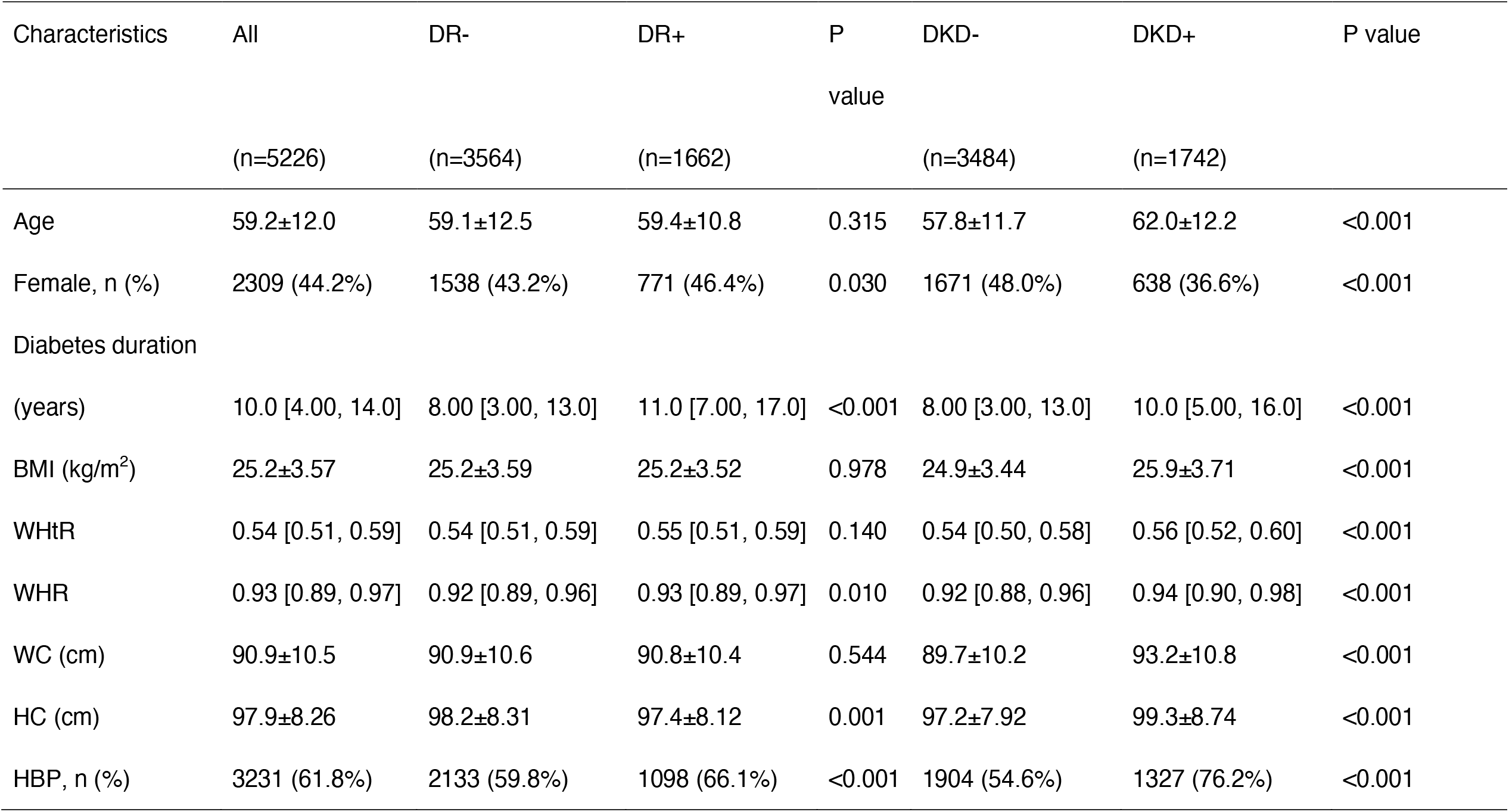
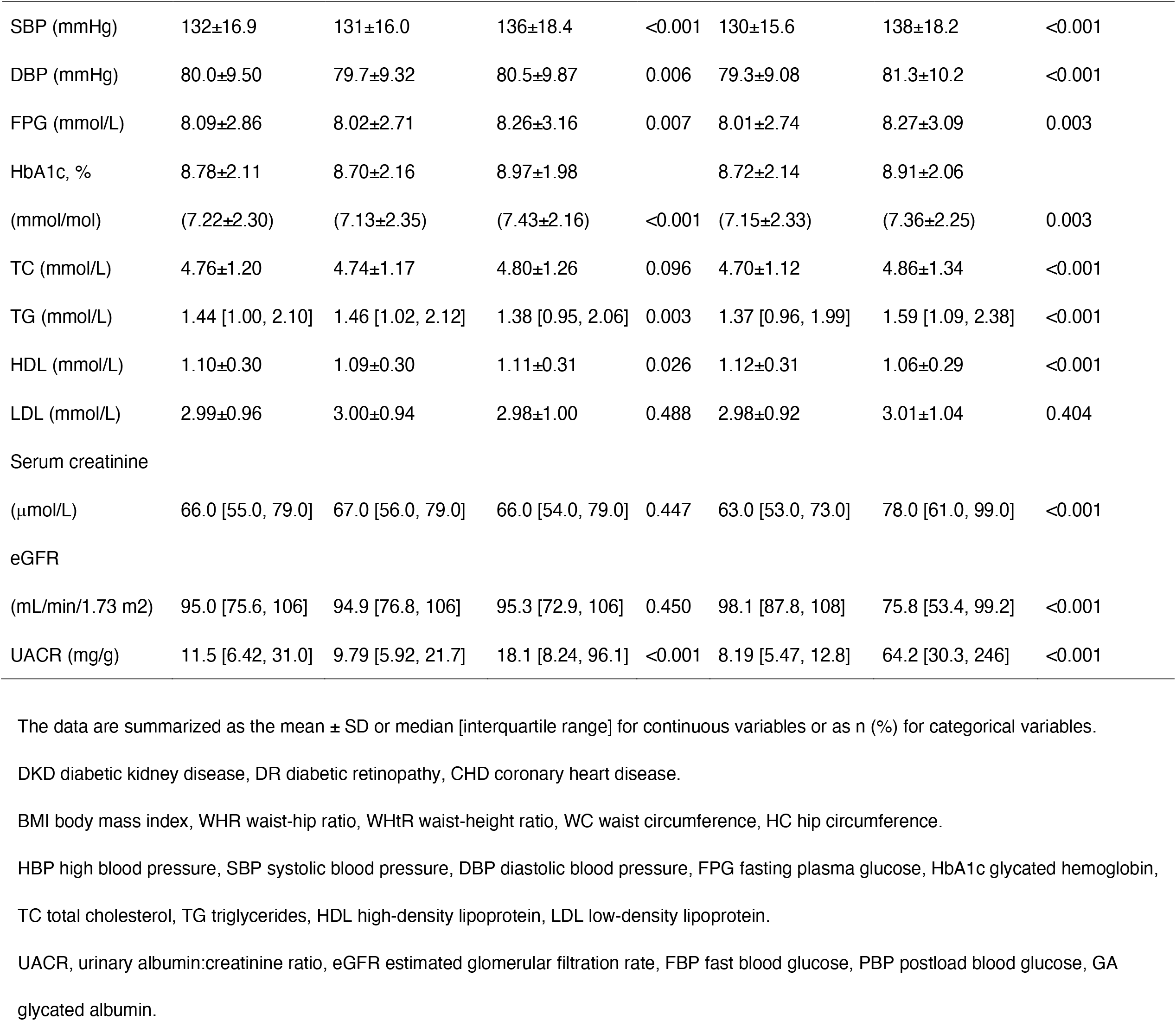
General characteristics of all participants with T2DM by DR and DKD.

| Characteristics | All | DR- | DR+ | P | DKD- | DKD+ | P value |
| --- | --- | --- | --- | --- | --- | --- | --- |
|  | (n=5226) | (n=3564) | (n=1662) | value | (n=3484) | (n=1742) |  |
| Age | 59.2±12.0 | 59.1±12.5 | 59.4±10.8 | 0.315 | 57.8±11.7 | 62.0±12.2 | <0.001 |
| Female, n (%) | 2309 (44.2%) | 1538 (43.2%) | 771 (46.4%) | 0.030 | 1671 (48.0%) | 638 (36.6%) | <0.001 |
| Diabetes duration |  |  |  |  |  |  |  |
| (years) | 10.0 [4.00, 14.0] | 8.00 [3.00, 13.0] | 11.0 [7.00, 17.0] | <0.001 | 8.00 [3.00, 13.0] | 10.0 [5.00, 16.0] | <0.001 |
| BMI (kg/m<sup>2</sup>) | 25.2±3.57 | 25.2±3.59 | 25.2±3.52 | 0.978 | 24.9±3.44 | 25.9±3.71 | <0.001 |
| WHtR | 0.54 [0.51, 0.59] | 0.54 [0.51, 0.59] | 0.55 [0.51, 0.59] | 0.140 | 0.54 [0.50, 0.58] | 0.56 [0.52, 0.60] | <0.001 |
| WHR | 0.93 [0.89, 0.97] | 0.92 [0.89, 0.96] | 0.93 [0.89, 0.97] | 0.010 | 0.92 [0.88, 0.96] | 0.94 [0.90, 0.98] | <0.001 |
| WC (cm) | 90.9±10.5 | 90.9±10.6 | 90.8±10.4 | 0.544 | 89.7±10.2 | 93.2±10.8 | <0.001 |
| HC (cm) | 97.9±8.26 | 98.2±8.31 | 97.4±8.12 | 0.001 | 97.2±7.92 | 99.3±8.74 | <0.001 |
| HBP, n (%) | 3231 (61.8%) | 2133 (59.8%) | 1098 (66.1%) | <0.001 | 1904 (54.6%) | 1327 (76.2%) | <0.001 |
| SBP (mmHg) | 132±16.9 | 131±16.0 | 136±18.4 | <0.001 | 130±15.6 | 138±18.2 | <0.001 |
| DBP (mmHg) | 80.0±9.50 | 79.7±9.32 | 80.5±9.87 | 0.006 | 79.3±9.08 | 81.3±10.2 | <0.001 |
| FPG (mmol/L) | 8.09±2.86 | 8.02±2.71 | 8.26±3.16 | 0.007 | 8.01±2.74 | 8.27±3.09 | 0.003 |
| HbA1c, % | 8.78±2.11 | 8.70±2.16 | 8.97±1.98 |  | 8.72±2.14 | 8.91±2.06 |  |
| (mmol/mol) | (7.22±2.30) | (7.13±2.35) | (7.43±2.16) | <0.001 | (7.15±2.33) | (7.36±2.25) | 0.003 |
| TC (mmol/L) | 4.76±1.20 | 4.74±1.17 | 4.80±1.26 | 0.096 | 4.70±1.12 | 4.86±1.34 | <0.001 |
| TG (mmol/L) | 1.44 [1.00, 2.10] | 1.46 [1.02, 2.12] | 1.38 [0.95, 2.06] | 0.003 | 1.37 [0.96, 1.99] | 1.59 [1.09, 2.38] | <0.001 |
| HDL (mmol/L) | 1.10±0.30 | 1.09±0.30 | 1.11±0.31 | 0.026 | 1.12±0.31 | 1.06±0.29 | <0.001 |
| LDL (mmol/L) | 2.99±0.96 | 3.00±0.94 | 2.98±1.00 | 0.488 | 2.98±0.92 | 3.01±1.04 | 0.404 |
| Serum creatinine |  |  |  |  |  |  |  |
| (μmol/L) | 66.0 [55.0, 79.0] | 67.0 [56.0, 79.0] | 66.0 [54.0, 79.0] | 0.447 | 63.0 [53.0, 73.0] | 78.0 [61.0, 99.0] | <0.001 |
| eGFR |  |  |  |  |  |  |  |
| (mL/min/1.73 m<sup>2</sup>) | 95.0 [75.6, 106] | 94.9 [76.8, 106] | 95.3 [72.9, 106] | 0.450 | 98.1 [87.8, 108] | 75.8 [53.4, 99.2] | <0.001 |
| UACR (mg/g) | 11.5 [6.42, 31.0] | 9.79 [5.92, 21.7] | 18.1 [8.24, 96.1] | <0.001 | 8.19 [5.47, 12.8] | 64.2 [30.3, 246] | <0.001 |

The data are summarized as the mean ± SD or median [interquartile range] for continuous variables or as n (%) for categorical variables.
DKD diabetic kidney disease, DR diabetic retinopathy, CHD coronary heart disease.
BMI body mass index, WHR waist-hip ratio, WHtR waist-height ratio, WC waist circumference, HC hip circumference.
HBP high blood pressure, SBP systolic blood pressure, DBP diastolic blood pressure, FPG fasting plasma glucose, HbA1c glycated hemoglobin,
TC total cholesterol, TG triglycerides, HDL high-density lipoprotein, LDL low-density lipoprotein.
UACR, urinary albumin:creatinine ratio, eGFR estimated glomerular filtration rate, FBP fast blood glucose, PBP postload blood glucose, GA glycated albumin.

### 3.2 Associations of anthropometric indices with DR and DKD

Table 2 and 3 show the associations of anthropometric indices with DR and DKD, which involved BMI, WHR, WHtR, WC and HC.

**Table 2.**
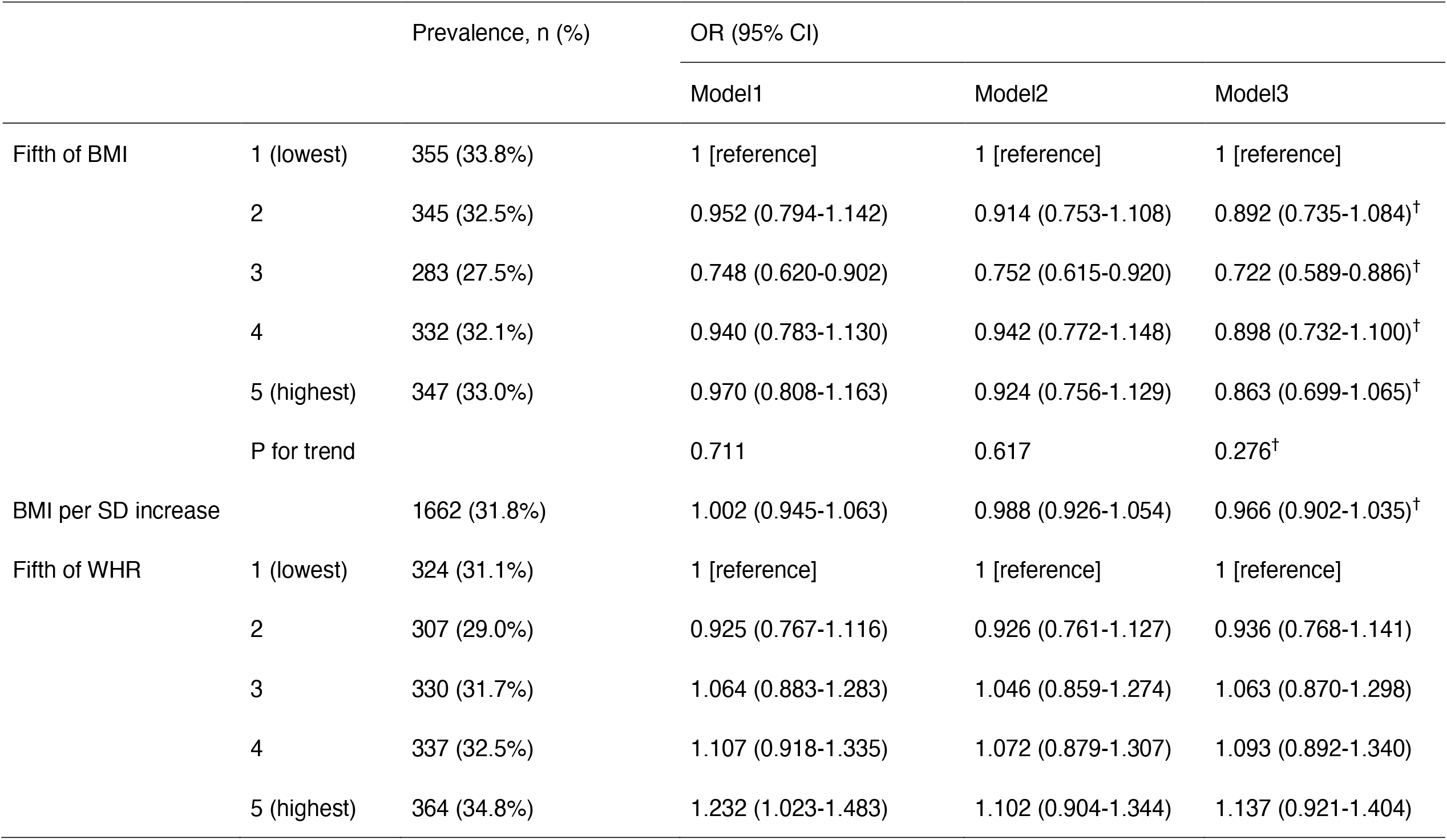
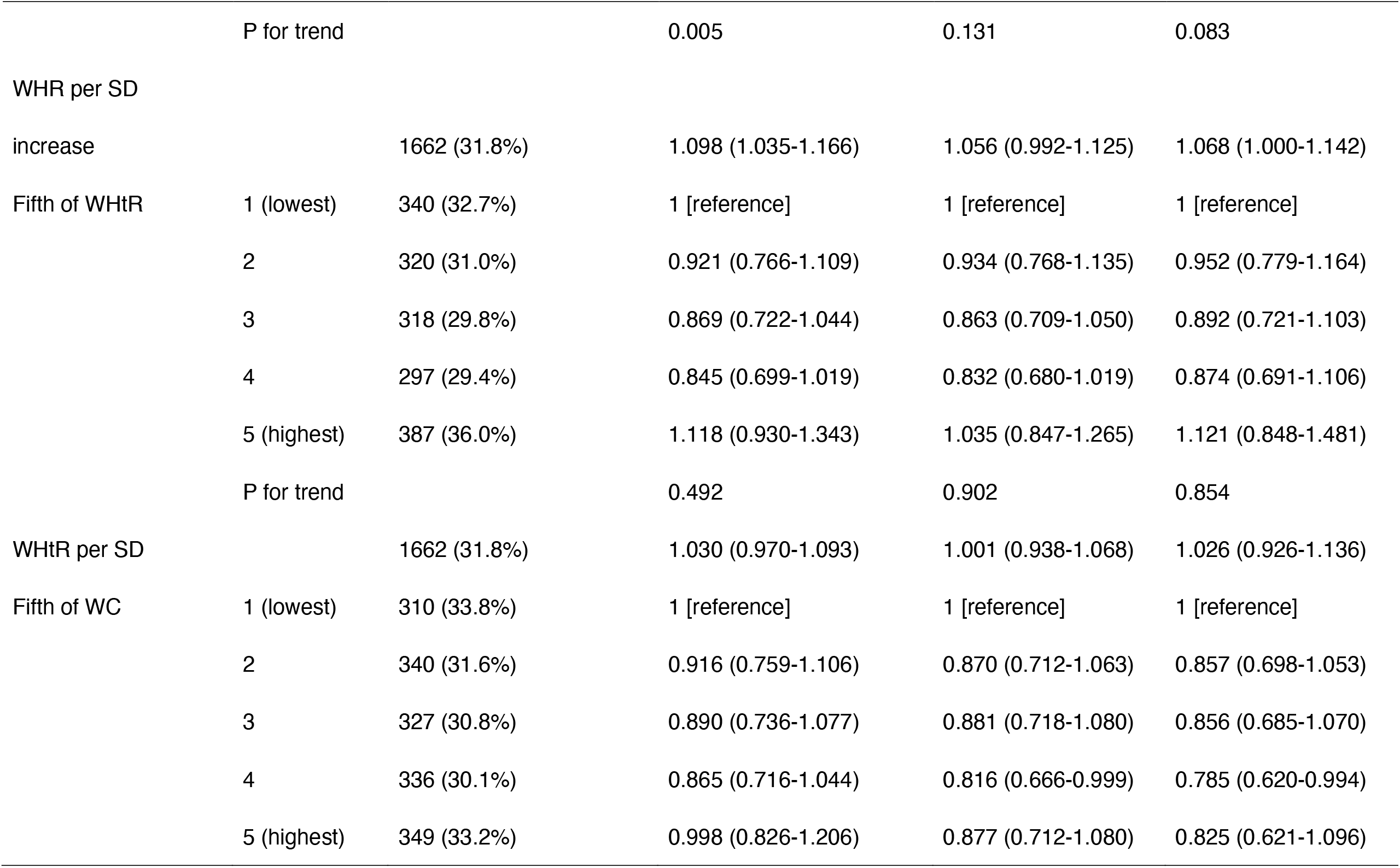
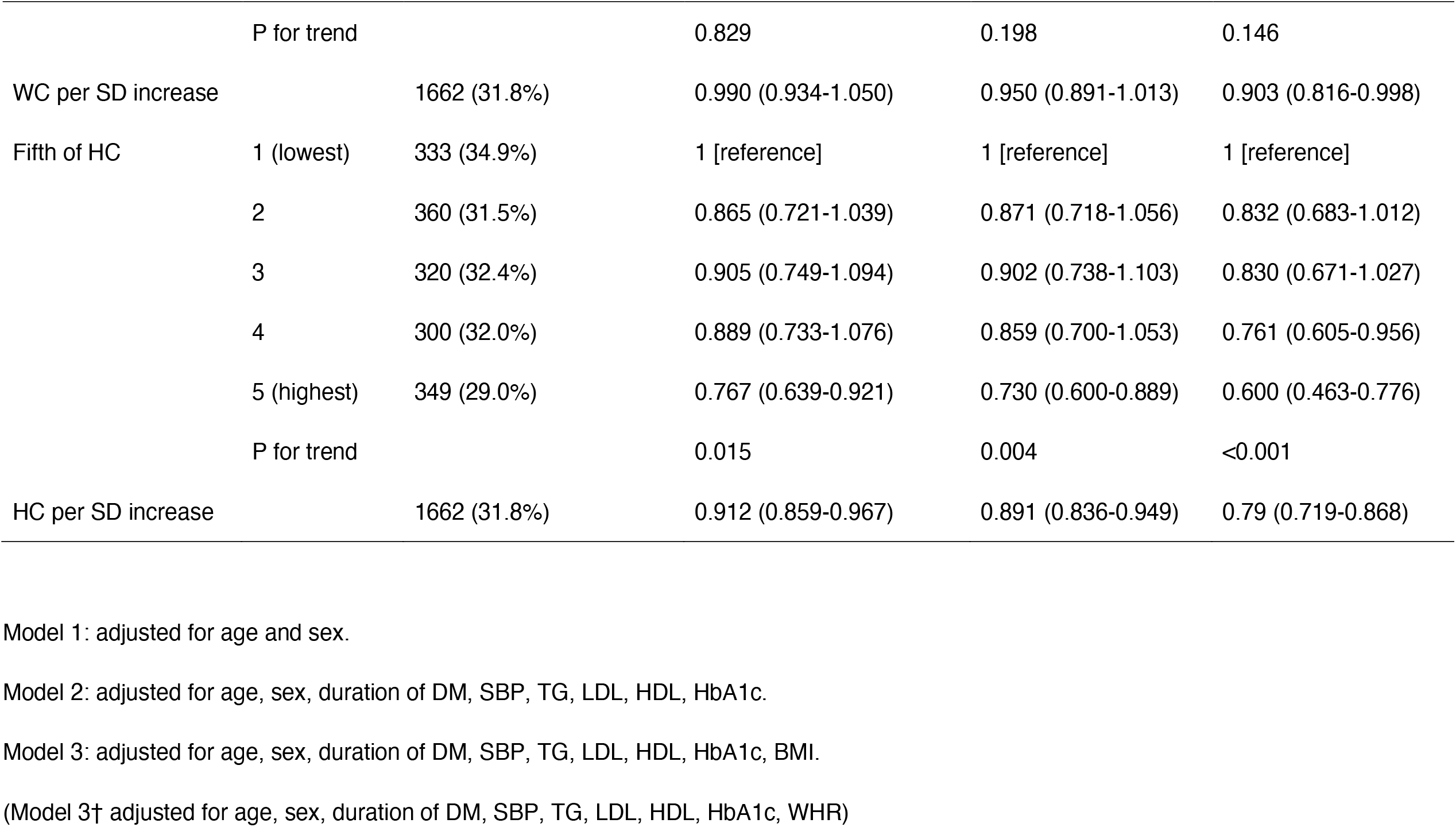
Odds ratios (95% CI) of DR according to anthropometric indices.

**Table 3.**
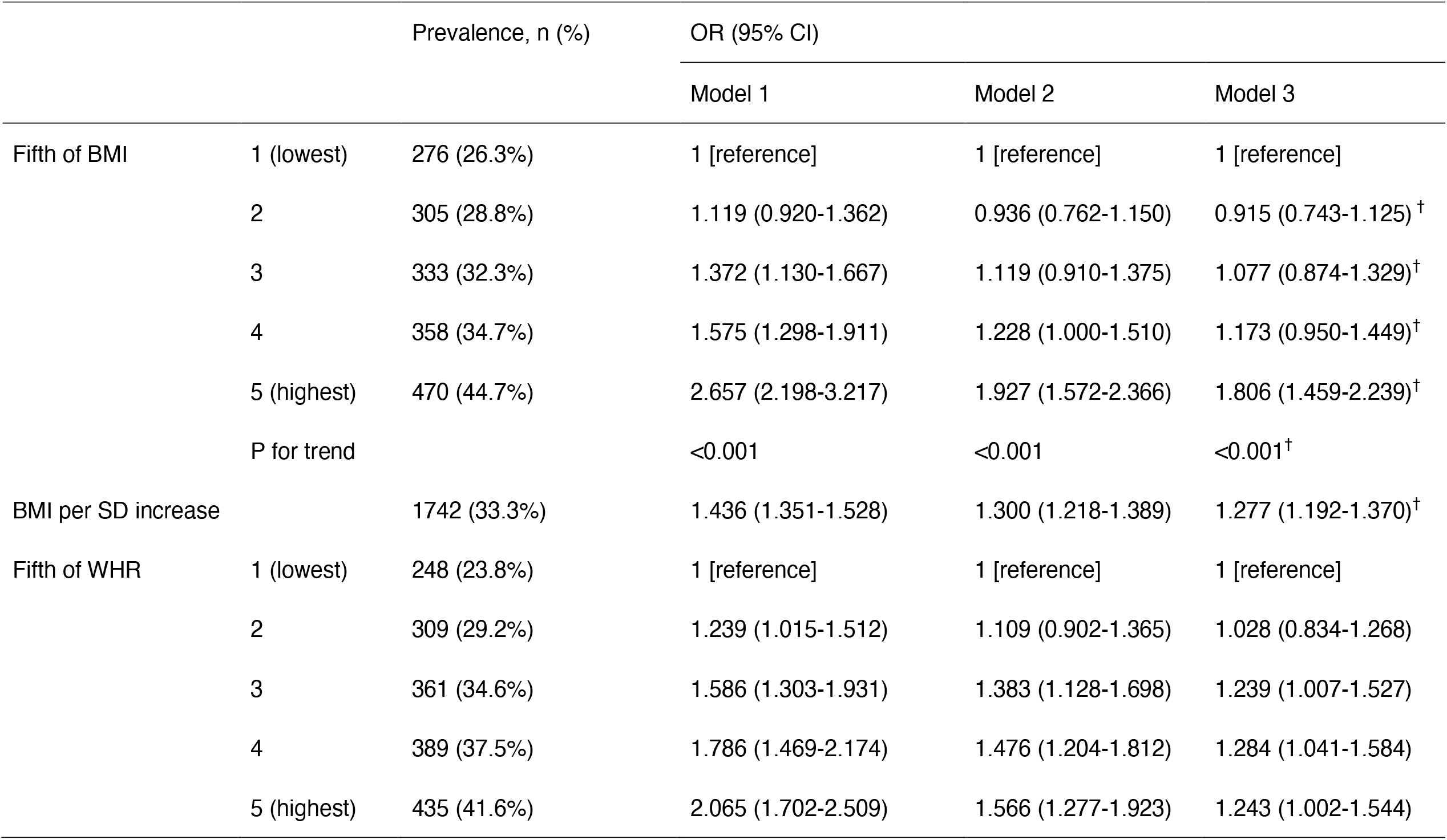
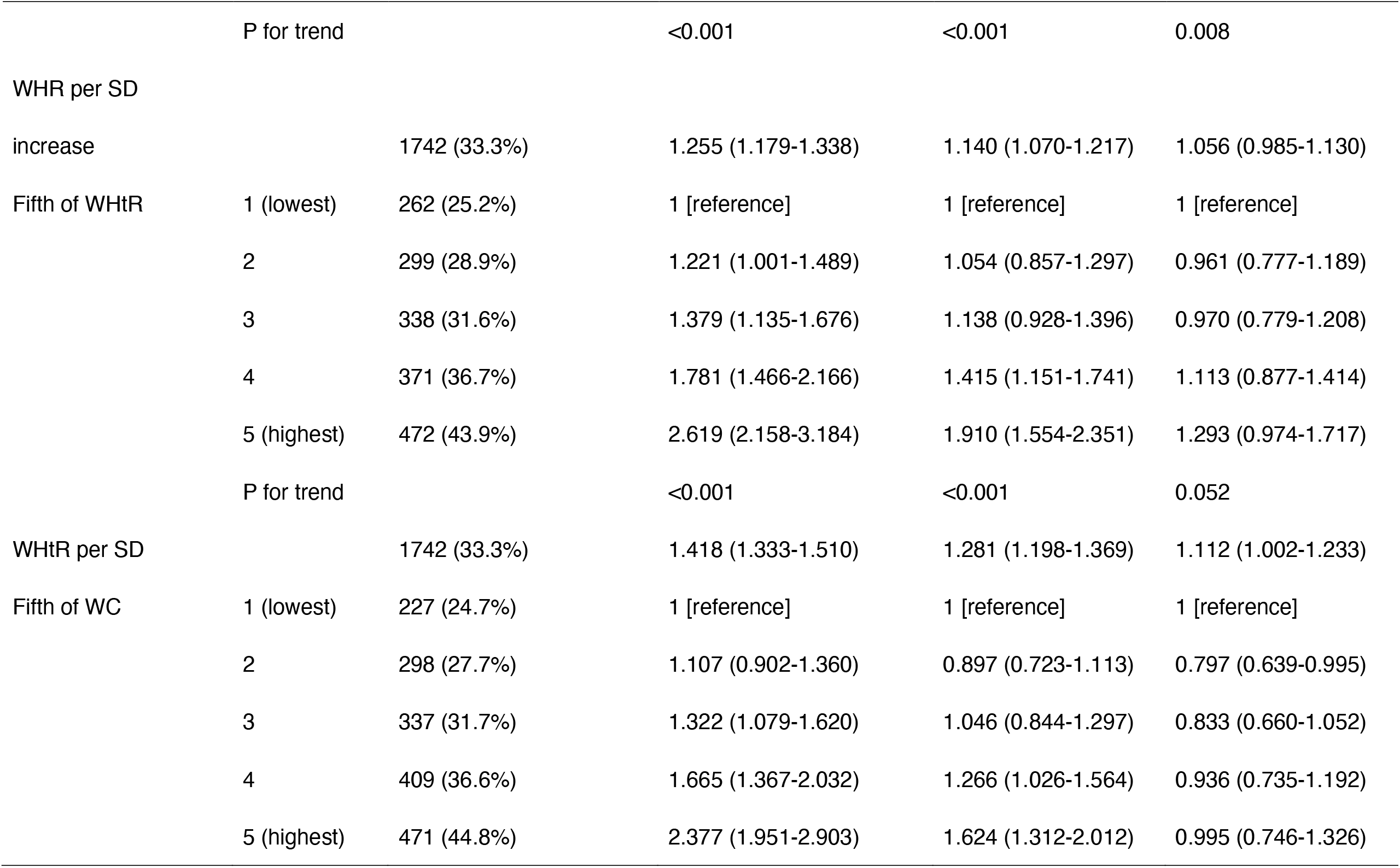
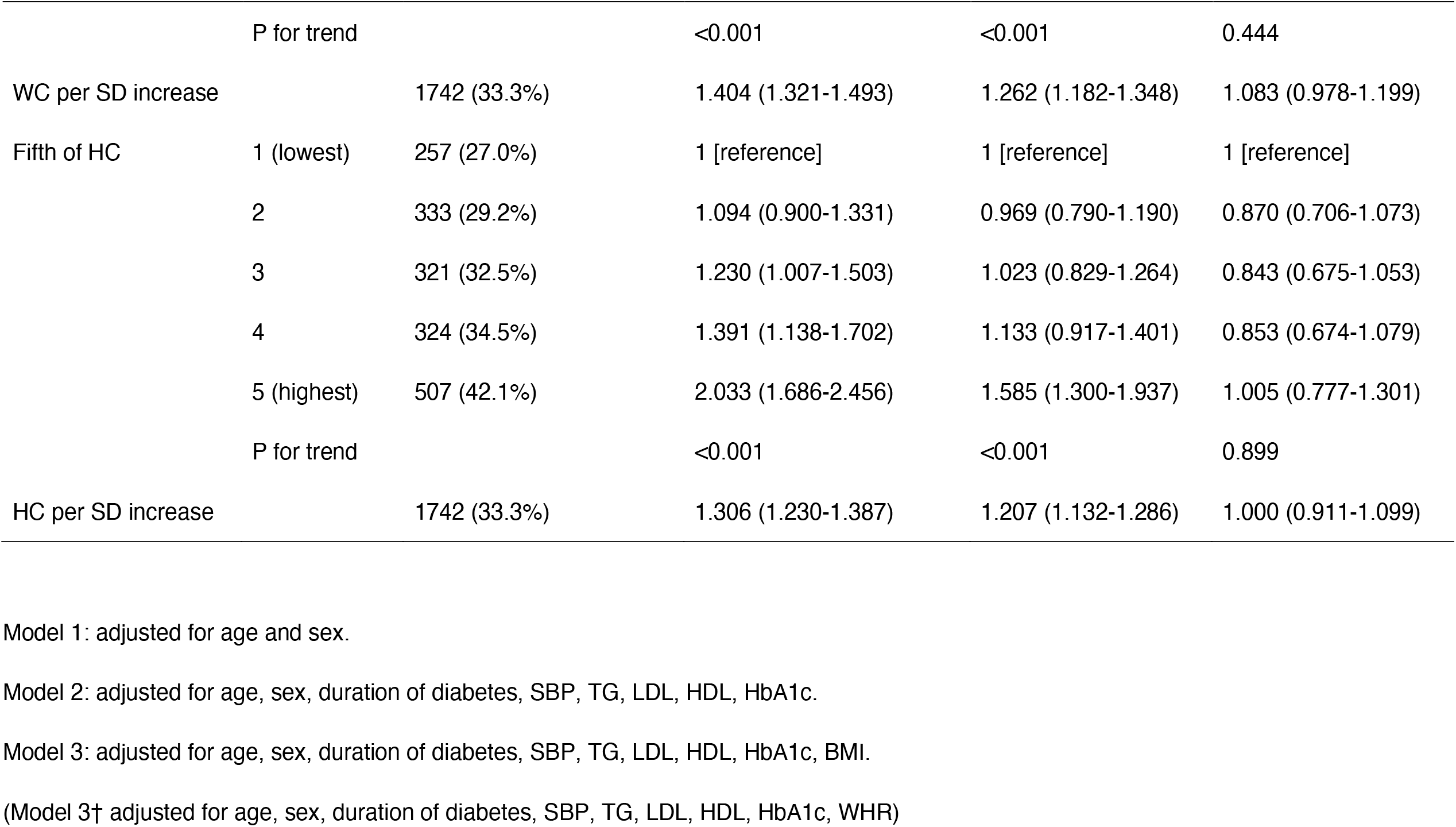
Odds ratio (95% CI) of DKD according to anthropometric indices.

HC showed an independent inverse association with DR. The OR based on the highest fifth of HC were 0.730 (0.600-0.889) in the multivariable model 2 adjusted for age, sex, duration of diabetes, SBP, TG, LDL, HDL and HbA1c, and 0.600 (0.463-0.776) in the model 3 further adjusted for BMI. Moreover, participants in the third fifth of BMI had the lowest OR for DR in the multivariable model 2 (OR 0.779, 95%CI 0.640-0.949) and model 3 further adjusted for WHR (OR 0.722, 95%CI 0.589-0.886).

In contrast, a positive association between all these anthropometric indices and DKD was shown in the multivariable model 2. In model 2, the OR based on the highest fifth of BMI, WHR, WHtR, WC and HC were 1.927 (1.572-2.366), 1.566 (1.277-1.923), 1.91 (1.554-2.351), 1.624 (1.312-2.012) and 1.585 (1.300-1.937) for DKD respectively. After further adjustment for BMI, the positive associations of WHR and WHtR with DKD were attenuated but still existed, while the positive associations of WC and HC disappeared. Besides, the positive association between BMI and DKD remained in the model 3 further adjusted for WHR.

In Figure1, we further used restricted cubic splines to flexibly model and visualize the relation of anthropometric indices with DR and DKD, after adjustment for age, sex, duration of diabetes, HbA1c, SBP, TG, LDL and HDL.

**Figure 1.**
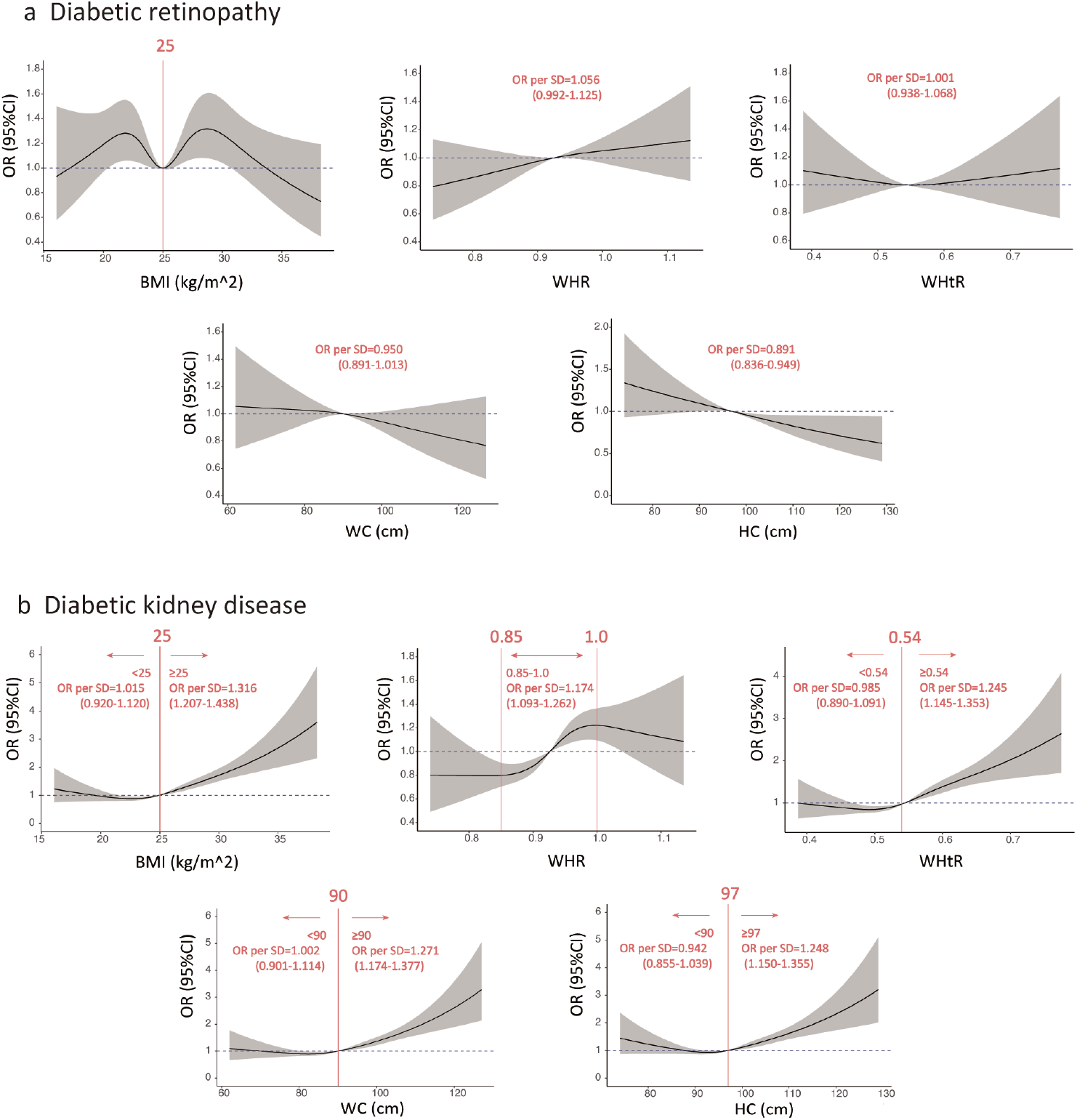
Associations between anthropometric indices and risk of DR and DKD. a Associations between anthropometric indices and risk of DR. In the analysis of DR, knots placed at 5th, 27.5th, 50th, 72.5th and 95th for BMI, 10th, 50th and 90th percentile for WHR, WHtR, WC and HC. Test for non-linearity: BMI, p=0.018; WHR, p=0.624; WHtR, p=0.928; WC, p=0.592; HC, p=0.853. b Associations between anthropometric indices and risk of DKD. In the analysis of DKD, knots placed at 5th, 35th, 65th and 95th for BMI, WHR, WHtR and HC, 10th, 50th and 90th percentile for WC. Test for non-linearity: BMI, p<0.001; WHR, p=0.045; WHtR, p=0.022; WC, p=0.002; HC, p<0.001. All the anthropometric indices were assessed as a continuous variable using restricted cubic spine regression, adjusted for age, sex, duration of diabetes, HbA1c, SBP, TG, LDL and HDL. Reference point was median value for each of anthropometric indices (25 for BMI, 0.93 for WHR, 0.54 for WHtR, 90 for WC, 97 for HC).

The analyses provided significant evidence of the non-linear associations between BMI and DR (p <0.05), while WHR, WHtR, WC and HC showed a significant linear relationship with DR (all p >0.05). The OR per SD higher of WHR, WHtR, WC and HC was 1.056 (0.992-1.125), 1.001(0.938-1.068), 0.950 (0.891-1.013) and 0.891 (0.836-0.949) respectively. Moreover, the OR of DR was relatively low at the median value of BMI.

We also found evidence of non-linear associations of all these anthropometric indices with DKD (all p <0.05). Beyond the threshold at about the median value of parameters, higher BMI, WHtR, WC and HC contribute significantly to DKD development. But there was no increased risk in the lower range of BMI, WHtR, WC and HC. Above the median value, the OR per SD higher of BMI, WHtR, WC and HC were 1.316 (1.207-1.438), 1.245 (1.145-1.353), 1.271 (1.174-1.377) and 1.248 (1.150-1.355) respectively. Besides, an increased risk was shown in the middle range of WHR. The OR per SD higher of WHR was 1.174 (1.093-1.262) between 0.85 and 1.0.

### 3.3 Associations of anthropometric indices with DR and DKD stratified by sex

We further examined how the associations of anthropometric indices changed by sex in Table 4, Table S3 and Table S4. And the baseline characteristics of the male and female participants were shown in table S5 and table S6 respectively.

**Table 4.**
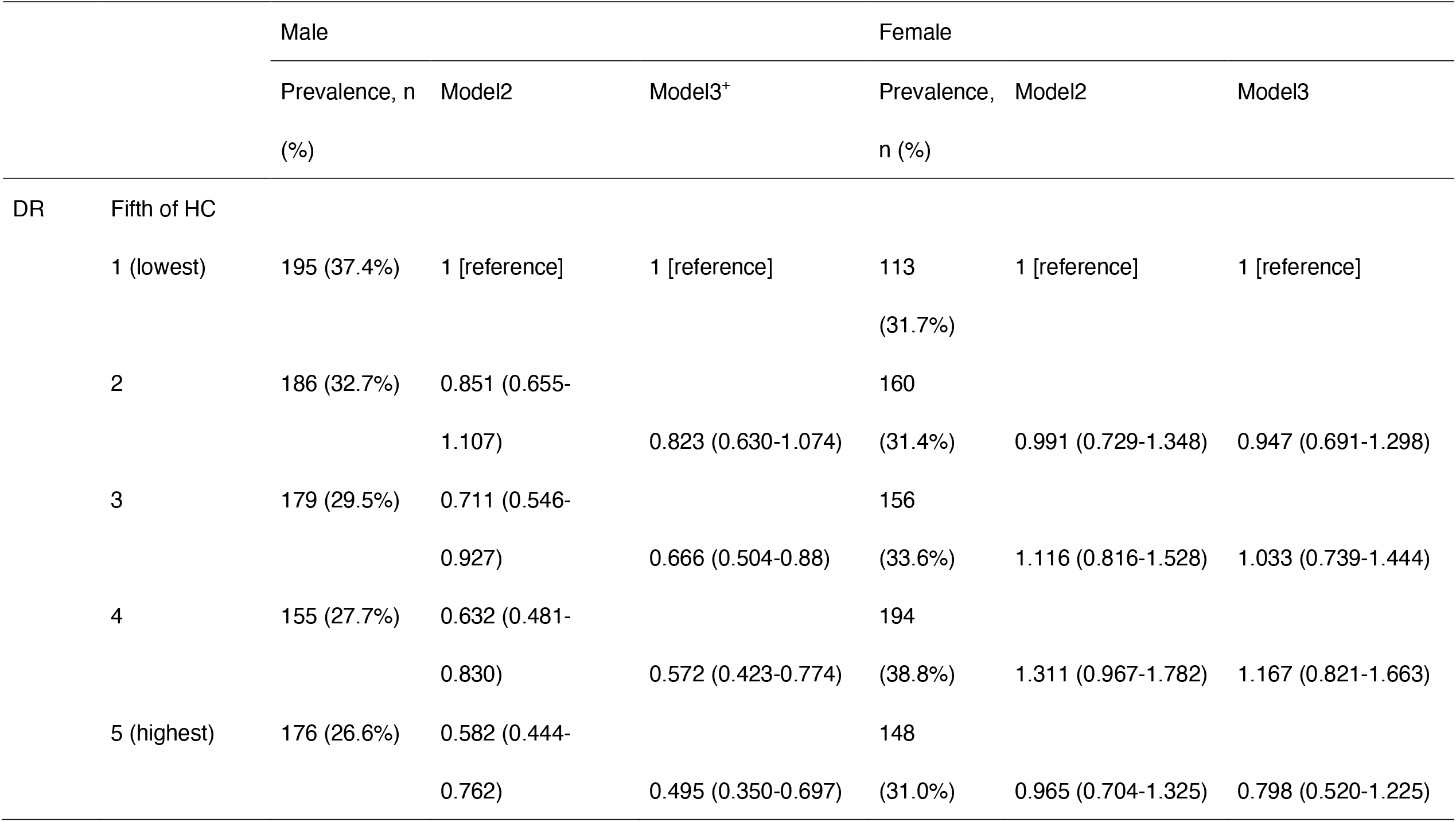
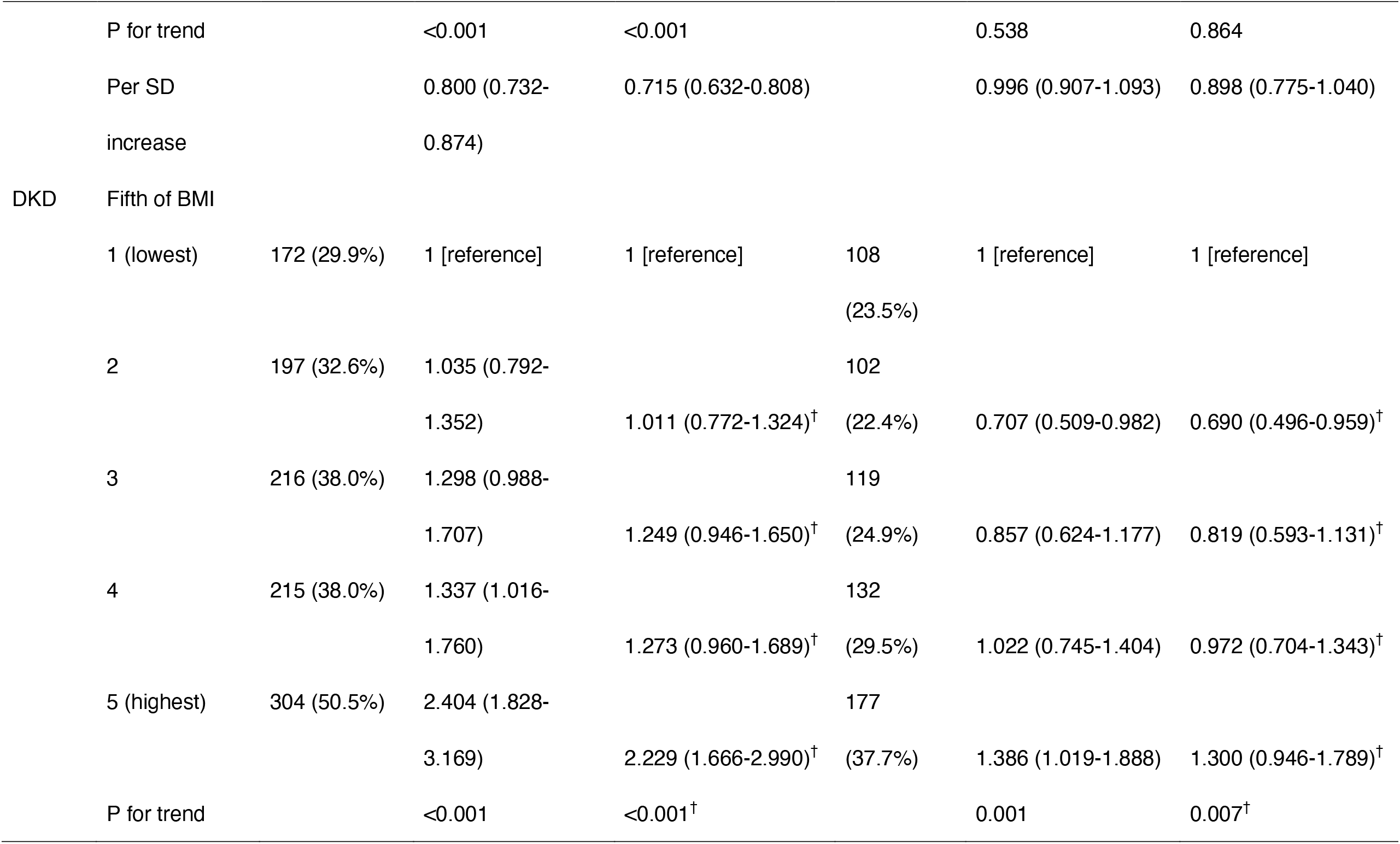
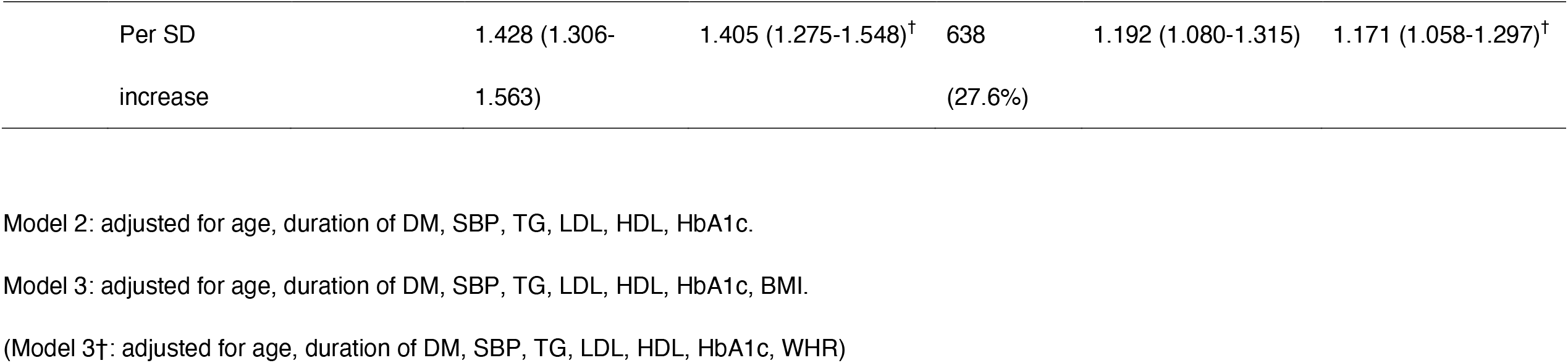
Odds ratios (95% CI) of DR and DKD according to BMI, WHR and HC stratified by sex.

The OR for DR based on the third fifth of BMI was the lowest across gender groups, although the OR in women was higher than that in men. We also found significant differences in WC (p=0.001 for interaction) and HC (p<0.001 for interaction) for DR across strata of sex (Table S3). The inverse associations of WC and HC with DR were observed in men, and still existed in model 3 further adjusted for BMI, whereas it was not found in women. The positive associations of anthropometric indices with DKD were generally consistent across gender groups. Further adjustment for WHR did not alter the significant positive association between BMI and DKD in men and women. Moreover, when further adjusted for BMI, the associations of WHR, WHtR, WC and HC with DKD were attenuated, among which only the positive association between WHtR and DKD in women remained significant.

We did several sensitivity analyses to verify our findings. The associations of BMI with DR and DKD remained robust with different cut points of BMI (Table S7). After mutually adjusting for WC and HC, HC was still inversely associated with DR, while the inverse association between WC and DR in men was attenuated (Table S8).

## 4. Discussion

In this cross-sectional study of Chinese participants with T2DM, we used logistic regression models and restricted cubic spine analysis to examine the different associations of anthropometric indices with DR and DKD. We found non-linear positive associations of all these anthropometric indices with DKD, while the associations of different anthropometric indices with DR varied dramatically. After taking WHR into account, BMI was still positively associated with DKD. In contrast, a BMI of around 25 kg/m^2^ was associated with the low risk of DR, and HC showed an independent and inverse association with DR in men.

To the best of our knowledge, our study is the first to analyze the non-linear associations of BMI, WHtR, WC and HC with DKD and DR using restricted cubic spline analysis in the Chinese population.

As is known, the associations between BMI and DR have been inconclusive. For example, in the Australian Diabetes Management Project ^11^, BMI showed a significantly positive association with DR, which was also reported in other western studies like the Netherlands Hoorn Study ^24^, whereas the Singapore Malay Eye Study (SiMES) reported an inverse association between BMI and DR using the quartiles of BMI ^25^. Besides, a meta-analysis of 27 clinical studies reported that compared with normal weight (BMI 18.5-24.9 kg/m^2^), neither overweight (BMI 25-29.9 kg/m^2^) nor obesity (BMI >=30 kg/m^2^) increased the risk of DR ^17^. Distinct with these findings, our research indicated that participants with a BMI of around the median value (25 kg/m^2^) have the lowest risk of DR, which added a new sight into the association between BMI and DR. The mechanism behind the association between BMI and DR is unclear. Too low BMI may reflect unintentional weight loss resulting from poorly controlled diabetic mellitus, which increases the risk of DR. And too high BMI may be associated with increased production of proinflammatory cytokines in adipocytes, leading to increased vascular permeability and a high risk of DR ^26^. However, due to the lack of cause and effect evidence in our cross-sectional study, more prospective studies focused on the relationship between BMI and DR are needed to verify our findings.

It has been established that a larger hip is associated with a lower risk of T2DM independently of WC and BMI according to previous studies ^27,28,29^. In our study, we found that HC also had a significant inverse association with DR independently of BMI and WC in men. To our knowledge, there has been only one other study that investigated the independent relationship of HC with DR: a cross-sectional study involving 1773 patients with T2DM in India found a significant negative correlation between HC and DR ^30^. However, this previous study did not take the interdependence of BMI, WC or WHR with HC into consideration, and it also did not perform any sex-stratified analysis. The mechanism underlying the negative association of HC on DR in men is yet to be explored, but there is evidence that may partly explain this association. A larger hip could reflect a greater muscle mass in the gluteal region ^29^. Skeletal muscle mass is the main target of insulin and one major site of insulin resistance. Besides, a larger hip also indicates increased femoral and gluteal fat mass, which is less sensitive to lipolytic stimuli ^31^ and has an inverse relationship with arterial stiffness ^32^. One reason for the gender difference in the associations of HC with DR may be the distinction in fat distribution between men and women: men tend to store more visceral fat leading to an apple-shaped body while premenopausal women tend to accumulate more fat in the subcutaneous regions leading to a pear-shaped body ^33^. Furthermore, the positive association between adiposity and low-grade systemic inflammation was stronger in women than in men ^34^, which might partly explain why the inverse association between HC and DR disappeared in women.

Our findings on the positive associations of anthropometric indices with DKD were broadly in line with the majority of previous studies. For instance, BMI has been identified as a risk factor for DKD development based on a systematic review and meta-analysis of 20 cohorts ^35^. Another meta-analysis involving 15 cross-sectional studies reported that WC, WHR and WHtR were associated with a greater risk of DKD ^18^. Our study also elaborated the approximately J-shaped associations of BMI, WHR, WHtR, WC and HC with DKD and the S-shaped association between WHR and DKD. J-shaped associations have been reported in numerous previous studies related to obesity, while S-shaped associations are rarely observed. For instance, one cohort study of 3.6 million adults in the UK found J-shaped associations of BMI with overall mortality and most specific causes of death ^36^. WC also showed a J-shaped association with total mortality in a 12-year prospective cohort study of US adults ^37^. Moreover, there are several studies suggesting J-shape associations between BMI and kidney diseases. A cross-sectional study in Norway suggested a J-shaped association between BMI and risk of CKD ^38^. Another cross-sectional study of the Southeast Asian population reported a J-shaped association between BMI and proteinuria ^39^. Because there are few researches on the non-linear associations of anthropometric indices with DKD, more studies are necessary to verify our findings.

Besides, our research indicated that BMI was positively associated with DKD independent of WHR. When further adjusted for BMI, the positive association of WHR and WHtR with DKD still existed but attenuated, while positive associations of WC and HC were not significant. In contrast to our findings, one cross-sectional study and another 5-year prospective study in China reported that WC was significantly associated with risk of DKD after adjustment for BMI, and BMI was not related to the risk of DKD in WHtR-adjusted model ^40^. The inconsistency between our research and previous studies may partly be due to the different covariates involved in the adjusted models, for the BMI-adjusted model and WHtR-adjusted model used in the previous studies did not include any other covariate. Further studies considering the mutually confounding effect between generalized and abdominal obesity are necessary to examine the associations of anthropometric indices with DKD.

Our study revealed remarkable differences between the association of anthropometric indices with DKD and that with DR, which might indicate the tissue-specific mechanisms underlying the pathogenesis of DR and DKD. Lipid metabolism has been proven to play different roles in the development of DR and DKD ^41^. Tissue-specific lipid alterations have been observed in mice with diabetes ^42^. Besides, analysis of metabolic flux revealed the difference of glucose and fatty acid metabolism in the progression of DR and DKD ^43^. Further researches investigating the difference between the pathogenesis of DR and DKD are warranted.

Our study has several strengths. Firstly, it is the first study to assess the non-linear associations of anthropometric indices with DR and DKD using restricted cubic spline concurrently. Secondly, the interdependence between general obesity and abdominal obesity was taken into account in our study. Thirdly, we did several sex-stratified and sensitivity analyses to verify the robustness of findings. In addition, we recruited a relatively large clinical sample of patients with T2DM and performed this study with a comprehensive and standardized clinical assessment protocol.

There are also some limitations in our study. Firstly, because of the cross-sectional nature of this study, causal inference of these anthropometric indices with DR and DKD cannot be determined. Secondly, our study was conducted in a hospital, which may affect the generalizability of findings. Lastly, because of the low numbers of participants with ESRD or proliferative diabetic retinopathy (PDR), we did not investigate the associations of anthropometric indices with ESRD and PDR.

In conclusion, this study revealed different associations of anthropometric indices with DR and DKD in Chinese patients with T2DM. A BMI of around 25 kg/m^2^ and a large hip might be related to the relatively low risk of DR, while a higher BMI, WHR, WHtR, WC or HC was associated with a higher risk of DKD. Our findings support recommendations to maintain a BMI of around 25 kg/m^2^, a low WHR, a low WHtR and a large hip for prevention of DR and DKD. More longitudinal researches are warranted to verify our findings.

## Data Availability

All data produced in the present study are available upon reasonable request to the authors.

## Acknowledgements

The authors thank the clinical staffs of diabetes and ophthalmology clinics of the Shanghai General Hospital and the participants in this study for their valuable contributions.

## Author Contributions

YJW, HBC and ZZ designed the research. XP, CFG, CXL and HBC collected the research data. YJW, XP and BL analyzed the data. YJW drafted the manuscript. ZZ, HBC and CDZ assumed primary responsibility for the final content. YJW and XP contributed equally to this work. All authors read and approved the final manuscript.

## Colors in Figures

#C15858 was used in Figure 1 (Figure1 can be printed without color).

