## Supplementary tables for "Different Associations of Anthropometric Indices with Diabetic Retinopathy and Diabetic Kidney Disease in Chinese Patients with Type 2 Diabetes Mellitus"

Table S1. Spearman correlations among anthropometric indices.

| | BMI | HC | WC | WCHC | WHtR |
|---|---|---|---|---|---|
| BMI | 1 | 0.755908062 | 0.776252179 | 0.365698709 | 0.757133368 |
| HC | 0.755908062 | 1 | 0.803403046 | 0.117141996 | 0.717727487 |
| WC | 0.776252179 | 0.803403046 | 1 | 0.680938749 | 0.900064397 |
| WCHC | 0.365698709 | 0.117141996 | 0.680938749 | 1 | 0.621562644 |
| WHtR | 0.757133368 | 0.717727487 | 0.900064397 | 0.621562644 | 1 |

BMI body mass index, WHR waist-hip ratio, WHtR waist-height ratio, WC waist circumference, HC hip circumference.

All correlations were significant $p<0.001$.

Table S2. Location of knots. From Harrell (2001), Regression Modeling Strategies.

| Number of knots K | Knots locations expressed in quantiles of the x variable | | | | |
|---|---|---|---|---|---|
| 3 | 0.1 | 0.5 | 0.9 | | |
| 4 | 0.05 | 0.35 | 0.65 | 0.95 | |
| 5 | 0.05 | 0.275 | 0.5 | 0.725 | 0.95 |

Table S3. Interaction tests between sex and anthropometric indices with the presence of DR and DKD.

| | | Test for interaction | |
|---|---|---|---|
| | | Model 2 | Model 3 |
| DR | BMI | 0.020 | 0.014 |
| | WHR | 0.454 | 0.497 |
| | WHtR | 0.010 | 0.010 |
| | WC | 0.001 | 0.001 |
| | HC | <0.001 | <0.001 |
| DKD | BMI | 0.016 | 0.022 |
| | WHR | 0.242 | 0.520 |
| | WHtR | 0.031 | 0.032 |
| | WC | 0.084 | 0.049 |
| | HC | 0.071 | 0.024 |

Model 2: adjusted for age, sex, duration of DM, SBP, TG, LDL, HDL, HbA1c.

Model 3: further including BMI for analyses of WHR, WHtR, WC and HC, or further including WHR for analyses of BMI.

Table S4. Odds ratios (95% CI) of DR and DKD according to BMI, WHR, WHtR, WC and HC stratified by sex.

| | | Male | | | Female | | |
|---|---|---|---|---|---|---|---|
| | | Prevalence, n (%) | Model2 | Model3 | Prevalence, n (%) | Model2 | Model3 |
| DR | Fifth of BMI | | | | | | |
| | 1 (lowest) | 203 (35.2%) | 1 [reference] | 1 [reference] | 147 (32.0%) | 1 [reference] | 1 [reference] |
| | 2 | 192 (31.8%) | 0.869 (0.672-1.125) | 0.844 (0.651-1.095)† | 149 (32.7%) | 0.910 (0.677-1.222) | 0.891 (0.662-1.199)† |
| | 3 | 147 (25.8%) | 0.656 (0.497-0.863) | 0.624 (0.471-0.826)† | 145 (30.4%) | 0.892 (0.664-1.198) | 0.857 (0.635-1.157)† |
| | 4 | 175 (30.9%) | 0.874 (0.666-1.145) | 0.822 (0.622-1.086)† | 153 (34.2%) | 1.035 (0.768-1.394) | 0.990 (0.731-1.342)† |
| | 5 (highest) | 174 (28.9%) | 0.747 (0.566-0.984) | 0.680 (0.505-0.913)† | 177 (37.7%) | 1.130 (0.841-1.519) | 1.068 (0.787-1.450)† |
| | P for trend | | 0.081 | 0.026† | | 0.248 | 0.447† |
| | Per SD increase | 891 (30.5%) | 0.928 (0.848-1.015) | 0.897 (0.812-0.989)† | 771 (33.4%) | 1.052 (0.957-1.155) | 1.032 (0.935-1.139)† |
| | Fifth of WHR | | | | | | |
| | 1 (lowest) | 191 (32.7%) | 1 [reference] | 1 [reference] | 140 (30.5%) | 1 [reference] | 1 [reference] |
| | 2 | 172 (29.6%) | 0.919 (0.709-1.191) | 0.983 (0.756-1.278) | 134 (28.7%) | 0.896 (0.665-1.206) | 0.888 (0.658-1.197) |
| | 3 | 152 (26.2%) | 0.741 (0.567-0.968) | 0.766 (0.585-1.002) | 149 (32.3%) | 1.038 (0.772-1.397) | 1.022 (0.756-1.382) |
| | 4 | 167 (30.0%) | 0.937 (0.718-1.221) | 0.994 (0.761-1.299) | 178 (38.0%) | 1.234 (0.923-1.652) | 1.214 (0.903-1.633) |
| | 5 (highest) | 209 (34.0%) | 0.990 (0.763-1.284) | 1.091 (0.838-1.421) | 170 (37.4%) | 1.187 (0.882-1.598) | 1.156 (0.848-1.577) |
| | P for trend | | 0.944 | 0.503 | | 0.046 | 0.077 |
| | Per SD increase | 891 (30.5%) | 1.041 (0.956-1.133) | 1.084 (0.989-1.189) | 771 (33.4%) | 1.077 (0.982-1.184) | 1.068 (0.970-1.178) |

| | | | | | | | |
|---|---|---|---|---|---|---|---|
| | Fifth of WHtR | | | | | | |
| | 1 (lowest) | 202 (34.5%) | 1 [reference] | 1 [reference] | 138 (29.9%) | 1 [reference] | 1 [reference] |
| | 2 | 167 (29.2%) | 0.826 (0.635-1.075) | 0.854 (0.650-1.120) | 130 (28.3%) | 0.855 (0.633-1.153) | 0.869 (0.637-1.183) |
| | 3 | 174 (29.3%) | 0.842 (0.646-1.096) | 0.888 (0.667-1.181) | 159 (34.6%) | 1.181 (0.878-1.590) | 1.214 (0.880-1.676) |
| | 4 | 170 (28.9%) | 0.805 (0.617-1.049) | 0.874 (0.639-1.195) | 180 (38.3%) | 1.369 (1.020-1.842) | 1.427 (1.006-2.029) |
| | 5 (highest) | 178 (30.7%) | 0.814 (0.620-1.069) | 0.927 (0.635-1.350) | 164 (35.7%) | 1.149 (0.850-1.553) | 1.225 (0.805-1.864) |
| | P for trend | | 0.166 | 0.742 | | 0.029 | 0.041 |
| | Per SD increase | | 0.931 (0.852-1.016) | 0.962 (0.839-1.103) | | 1.080 (0.982-1.188) | 1.098 (0.945-1.274) |
| | Fifth of WC | | | | | | |
| | 1 (lowest) | 196 (35.9%) | 1 [reference] | 1 [reference] | 116 (30.5%) | 1 [reference] | 1 [reference] |
| | 2 | 191 (32.0%) | 0.876 (0.674-1.138) | 0.868 (0.663-1.136) | 162 (32.1%) | 1.020 (0.753-1.385) | 1.022 (0.748-1.398) |
| | 3 | 152 (26.9%) | 0.679 (0.515-0.893) | 0.668 (0.495-0.899) | 155 (31.6%) | 1.058 (0.776-1.443) | 1.061 (0.759-1.486) |
| | 4 | 156 (27.9%) | 0.688 (0.522-0.907) | 0.673 (0.491-0.923) | 144 (33.8%) | 1.051 (0.764-1.448) | 1.055 (0.732-1.524) |
| | 5 (highest) | 196 (30.2%) | 0.715 (0.544-0.938) | 0.689 (0.476-0.997) | 194 (38.1%) | 1.239 (0.913-1.686) | 1.247 (0.824-1.892) |
| | P for trend | | 0.005 | 0.019 | | 0.152 | 0.329 |
| | Per SD increase | | 0.865 (0.791-0.945) | 0.809 (0.705-0.928) | | 1.054 (0.959-1.157) | 1.034 (0.892-1.196) |
| DKD | Fifth of WHR | | | | | | |
| | 1 (lowest) | 175 (30.0%) | 1 [reference] | 1 [reference] | 95 (20.7%) | 1 [reference] | 1 [reference] |
| | 2 | 199 (34.2%) | 1.180 (0.907-1.538) | 1.045 (0.799-1.367) | 107 (22.9%) | 0.999 (0.722-1.385) | 0.958 (0.691-1.330) |

| | | | | | | |
|---|---|---|---|---|---|---|
| 3 | 224 (38.6%) | 1.373 (1.056-1.787) | 1.192 (0.912-1.560) | 131 (28.4%) | 1.206 (0.878-1.662) | 1.112 (0.805-1.540) |
| 4 | 230 (41.4%) | 1.592 (1.223-2.076) | 1.275 (0.970-1.678) | 150 (32.1%) | 1.406 (1.029-1.926) | 1.295 (0.943-1.783) |
| 5 (highest) | 276 (44.9%) | 1.607 (1.238-2.089) | 1.119 (0.843-1.486) | 155 (34.1%) | 1.383 (1.008-1.903) | 1.211 (0.871-1.688) |
| P for trend | | <0.001 | 0.200 | | 0.005 | 0.061 |
| Per SD increase | 1104 (37.8%) | 1.178 (1.083-1.281) | 1.041 (0.950-1.141) | 638 (27.6%) | 1.111 (1.009-1.226) | 1.067 (0.962-1.180) |
| Fifth of WHtR | | | | | | |
| 1 (lowest) | 169 (28.8%) | 1 [reference] | 1 [reference] | 88 (19.1%) | 1 [reference] | 1 [reference] |
| 2 | 187 (32.7%) | 1.045 (0.798-1.369) | 0.888 (0.672-1.174) | 98 (21.3%) | 0.931 (0.665-1.304) | 0.919 (0.651-1.297) |
| 3 | 196 (33.1%) | 1.013 (0.775-1.326) | 0.774 (0.579-1.033) | 121 (26.4%) | 1.112 (0.801-1.546) | 1.087 (0.765-1.547) |
| 4 | 246 (41.8%) | 1.478 (1.134-1.928) | 0.973 (0.712-1.330) | 153 (32.6%) | 1.443 (1.049-1.992) | 1.394 (0.960-2.030) |
| 5 (highest) | 306 (52.8%) | 1.986 (1.518-2.602) | 1.033 (0.713-1.496) | 178 (38.8%) | 1.753 (1.277-2.415) | 1.660(1.073-2.575) |
| P for trend | | <0.001 | 0.716 | | <0.001 | 0.004 |
| Per SD increase | | 1.340 (1.229-1.462) | 1.070 (0.936-1.224) | | 1.230 (1.113-1.36) | 1.188 (1.015-1.388) |
| Fifth of WC | | | | | | |
| 1 (lowest) | 163 (29.9%) | 1 [reference] | 1 [reference] | 79 (20.8%) | 1 [reference] | 1 [reference] |
| 2 | 190 (31.9%) | 0.970 (0.738-1.274) | 0.825 (0.623-1.093) | 109 (21.6%) | 0.852 (0.606-1.202) | 0.812 (0.573-1.154) |
| 3 | 195 (34.5%) | 1.086 (0.823-1.434) | 0.807 (0.598-1.090) | 138 (28.2%) | 1.130 (0.809-1.583) | 1.031 (0.719-1.482) |
| 4 | 234 (41.8%) | 1.469 (1.117-1.935) | 0.984 (0.719-1.347) | 119 (27.9%) | 1.025 (0.725-1.451) | 0.904 (0.611-1.340) |
| 5 (highest) | 322 (49.5%) | 1.887 (1.441-2.477) | 0.982 (0.681-1.415) | 193 (37.9%) | 1.545 (1.118-2.147) | 1.269 (0.821-1.966) |
| P for trend | | <0.001 | 0.589 | | 0.001 | 0.180 |

| | | | | | | |
|---|---|---|---|---|---|---|
| Per SD increase | | 1.343 (1.231-1.466) | 1.075 (0.94-1.228) | | 1.187 (1.075-1.31) | 1.091 (0.934-1.272) |
| Fifth of HC | | | | | | |
| 1 (lowest) | 163 (31.3%) | 1 [reference] | 1 [reference] | 88 (24.6%) | 1 [reference] | 1 [reference] |
| 2 | 186 (32.7%) | 0.999 (0.761-1.314) | 0.872 (0.660-1.153) | 123 (24.1%) | 0.854 (0.612-1.194) | 0.789 (0.561-1.111) |
| 3 | 222 (36.6%) | 1.085 (0.829-1.422) | 0.841 (0.633-1.117) | 112 (24.1%) | 0.801 (0.570-1.128) | 0.701 (0.488-1.007) |
| 4 | 215 (38.4%) | 1.211 (0.921-1.593) | 0.818 (0.604-1.108) | 145 (29.0%) | 0.970 (0.699-1.349) | 0.793 (0.545-1.156) |
| 5 (highest) | 318 (48.1%) | 1.753 (1.346-2.287) | 0.931 (0.665-1.303) | 170 (35.6%) | 1.340 (0.969-1.861) | 0.969 (0.625-1.505) |
| P for trend | | <0.001 | 0.634 | | 0.015 | 0.992 |
| Per SD increase | | 1.275 (1.172-1.388) | 1.013 (0.900-1.140) | | 1.137 (1.031-1.254) | 0.985 (0.844-1.149) |

Model 2: adjusted for age, duration of DM, SBP, TG, LDL, HDL, HbA1c.

Model 3: adjusted for age, duration of DM, SBP, TG, LDL, HDL, HbA1c, BMI.

(Model 3† adjusted for age, duration of DM, SBP, TG, LDL, HDL, HbA1c, WHR)

Table S5. General characteristics of all male participants with T2DM by DR and DKD.

| Characteristics | All (n=2917) | DR- (n=2026) | DR+ (n=891) | P value | DKD- (n=1813) | DKD+ (n=1104) | P value |
|---|---|---|---|---|---|---|---|
| Age | 57.7±12.4 | 57.5±13.0 | 57.9±10.9 | 0.384 | 55.4±11.8 | 61.3±12.5 | <0.001 |
| Diabetes duration (years) | 8.00 [3.00, 13.0] | 7.00 [2.00, 12.0] | 10.0 [6.00, 16.0] | <0.001 | 7.00 [3.00, 12.0] | 10.0 [5.00, 15.0] | <0.001 |
| BMI (kg/m$^2$) | 25.2±3.30 | 25.2±3.59 | 25.2±3.52 | 0.052 | 24.8±3.14 | 25.8±3.44 | <0.001 |
| WHtR | 0.53 [0.50, 0.57] | 0.54 [0.50, 0.57] | 0.53 [0.50, 0.57] | 0.131 | 0.53 [0.49, 0.56] | 0.55 [0.51, 0.59] | <0.001 |
| WHR | 0.94 [0.90, 0.97] | 0.94 [0.90, 0.97] | 0.94 [0.90, 0.98] | 0.348 | 0.93 [0.89, 0.97] | 0.95 [0.91, 0.98] | <0.001 |
| WC (cm) | 92.1±9.99 | 92.4±9.97 | 91.3±9.97 | 0.006 | 90.8±9.46 | 94.1±10.5 | <0.001 |
| HC (cm) | 98.2±7.76 | 98.6±7.76 | 97.1±7.65 | <0.001 | 97.4±7.31 | 99.5±8.27 | <0.001 |
| HBP, n (%) | 1728 (59.2%) | 1171 (57.8%) | 557 (62.5%) | 0.019 | 913 (50.4%) | 815 (73.8%) | <0.001 |
| SBP (mmHg) | 131±16.7 | 130±15.7 | 134±18.3 | <0.001 | 128±15.0 | 136±18.0 | <0.001 |
| DBP (mmHg) | 80.4±9.54 | 80.2±9.45 | 81.0±9.70 | 0.026 | 79.8±9.14 | 81.5±10.1 | <0.001 |
| FBG (mmol/L) | 8.22±3.01 | 8.14±2.84 | 8.38±3.37 | 0.065 | 8.17±2.88 | 8.29±3.22 | 0.336 |
| HbA1c (%) | 8.87±2.17 | 8.81±2.23 | 8.99±2.04 | 0.032 | 8.87±2.22 | 8.87±2.09 | 0.975 |
| TC (mmol/L) | 4.60±1.18 | 4.59±1.18 | 4.61±1.19 | 0.669 | 4.53±1.10 | 4.72±1.29 | <0.001 |
| TG (mmol/L) | 1.40 [0.97, 2.09] | 1.44 [0.99, 2.12] | 1.35 [0.92, 2.01] | 0.003 | 1.35 [0.93, 1.98] | 1.52 [1.05, 2.31] | <0.001 |
| HDL (mmol/L) | 1.03±0.27 | 1.02±0.27 | 1.05±0.29 | 0.011 | 1.04±0.27 | 1.02±0.28 | 0.03 |
| LDL (mmol/L) | 2.91±0.94 | 2.92±0.91 | 2.89±0.98 | 0.378 | 2.89±0.88 | 2.94±1.02 | 0.194 |
| Serum creatinine(μmol/L) | 74.0 [65.0, 86.0] | 75.0 [66.0, 85.0] | 73.0 [64.0, 88.0] | 0.445 | 70.0 [63.0, 78.0] | 88.0 [72.0, 105] | <0.001 |
| Uric acid (μmol/L) | 345±91.7 | 347±93.4 | 341±87.5 | 0.094 | 327±81.6 | 374±99.4 | <0.001 |

| | | | | | | | |
|---|---|---|---|---|---|---|---|
| eGFR (mL/min/1.73 m2) | 90.1 [65.4, 101] | 89.7 [66.5, 100] | 90.9 [63.0, 101] | 0.639 | 94.3 [83.7, 103] | 61.3 [49.2, 93.1] | <0.001 |
| UACR (mg/g) | 10.4 [5.61, 30.7] | 8.81 [5.23, 21.9] | 17.6 [7.23, 110] | <0.001 | 6.92 [4.86, 11.4] | 60.2 [23.5, 256] | <0.001 |

Table S6. General characteristics of all female participants with T2DM by DR and DKD.

| Characteristics | All (n=2309) | DR- (n=1538) | DR+ (n=771) | P value | DKD- (n=1671) | DKD+ (n=638) | P value |
|---|---|---|---|---|---|---|---|
| Age | 61.1±11.2 | 61.1±11.6 | 61.1±10.4 | 0.97 | 60.3±11.0 | 63.0±11.4 | <0.001 |
| Diabetes duration (years) | 10.0 [5.00, 15.0] | 9.00 [4.00, 14.0] | 12.0 [8.00, 18.0] | <0.001 | 10.0 [4.00, 15.0] | 11.0 [6.00, 17.0] | <0.001 |
| BMI (kg/m$^2$) | 25.2±3.88 | 25.1±3.92 | 25.4±3.80 | 0.068 | 25.0±3.73 | 26.0±4.15 | <0.001 |
| WHtR | 0.56 [0.52, 0.61] | 0.56 [0.51, 0.61] | 0.57 [0.52, 0.61] | 0.003 | 0.55 [0.51, 0.60] | 0.58 [0.53, 0.62] | <0.001 |
| WHR | 0.91 [0.87, 0.96] | 0.91 [0.87, 0.96] | 0.92 [0.88, 0.96] | 0.001 | 0.91 [0.87, 0.95] | 0.92 [0.89, 0.97] | <0.001 |
| WC (cm) | 89.4±11.0 | 89.0±11.1 | 90.1±10.9 | 0.024 | 88.5±10.8 | 91.7±11.3 | <0.001 |
| HC (cm) | 97.6±8.84 | 97.5±8.94 | 97.7±8.62 | 0.706 | 97.1±8.52 | 98.9±9.49 | <0.001 |
| HBP, n (%) | 1503 (65.1%) | 962 (62.5%) | 541 (70.2%) | <0.001 | 991 (59.3%) | 512 (80.3%) | <0.001 |
| SBP (mmHg) | 134±17.1 | 132±16.2 | 137±18.4 | <0.001 | 132±16.0 | 140±18.2 | <0.001 |
| DBP (mmHg) | 79.4±9.43 | 79.1±9.10 | 79.9±10.0 | 0.073 | 78.7±8.99 | 81.0±10.3 | <0.001 |
| FBG (mmol/L) | 7.94±2.65 | 7.85±2.52 | 8.12±2.89 | 0.032 | 7.83±2.57 | 8.24±2.85 | 0.002 |
| HbA1c, % (mmol/mol) | 8.68±2.03 (7.11±2.21) | 8.54±2.07 (6.96±2.26) | 8.95±1.92 (7.41±2.09) | <0.001 | 8.57±2.03 (6.99±2.21) | 8.97±1.99 (7.43±2.17) | <0.001 |
| TC (mmol/L) | 4.96±1.19 | 4.93±1.13 | 5.01±1.31 | 0.126 | 4.89±1.11 | 5.12±1.38 | <0.001 |
| TG (mmol/L) | 1.47 [1.03, 2.12] | 1.47 [1.05, 2.12] | 1.47 [0.99, 2.12] | 0.346 | 1.40 [0.99, 2.00] | 1.69 [1.19, 2.49] | <0.001 |
| HDL (mmol/L) | 1.18±0.32 | 1.18±0.32 | 1.18±0.32 | 0.967 | 1.21±0.32 | 1.13±0.30 | <0.001 |
| LDL (mmol/L) | 3.09±0.98 | 3.09±0.98 | 3.08±1.00 | 0.715 | 3.08±0.95 | 3.12±1.07 | 0.399 |
| Serum creatinine(μmol/L) | 56.0 [48.0, 65.0] | 56.0 [48.0, 64.0] | 56.0 [48.0, 67.0] | 0.286 | 55.0 [48.0, 63.0] | 60.0 [50.0, 79.0] | <0.001 |

| | | | | | | | |
|---|---|---|---|---|---|---|---|
| eGFR | | | | | | | |
| (mL/min/1.73 m2) | 101 [88.3, 111] | 102 [89.4, 111] | 100 [86.0, 111] | 0.068 | 102 [92.7, 111] | 95.8 [70.7, 108] | <0.001 |
| UACR (mg/g) | 12.8 [7.71, 31.3] | 10.9 [7.32, 21.6] | 18.4 [9.29, 75.7] | <0.001 | 9.39 [6.79, 14.2] | 71.7 [37.0, 239] | <0.001 |

Table S7. Sensitivity analysis of BMI in relation to DR and DKD.

| | N | DR | | DKD | |
|---|---|---|---|---|---|
| | | Model 1 | Model 2 | Model 1 | Model 2 |
| BMI strata | | | | | |
| <19.5 | 198 | 1.017 (0.709-1.446) | 1.067 (0.726-1.556) | 0.715 (0.488-1.033) | 1.109 (0.743-1.637) |
| 19.5-20.5 | 195 | 1.246 (0.877-1.759) | 1.192 (0.823-1.713) | 0.676 (0.460-0.979) | 0.852 (0.573-1.254) |
| 20.5-21.5 | 311 | 1.681 (1.260-2.240) | 1.640 (1.210-2.222) | 0.944 (0.696-1.275) | 1.108 (0.807-1.518) |
| 21.5-22.5 | 436 | 1.156 (0.883-1.512) | 1.105 (0.834-1.463) | 0.705 (0.530-0.934) | 0.799 (0.595-1.069) |
| 22.5-23.5 | 557 | 1.246 (0.971-1.600) | 1.165 (0.898-1.513) | 0.850 (0.657-1.098) | 0.887 (0.680-1.157) |
| 23.5-24.5 | 651 | 1.206 (0.948-1.536) | 1.178 (0.916-1.516) | 0.892 (0.699-1.138) | 0.928 (0.721-1.194) |
| 24.5-25.5 | 628 | 1 [reference] | 1 [reference] | 1 [reference] | 1 [reference] |
| 25.5-26.5 | 572 | 1.174 (0.915-1.507) | 1.163 (0.897-1.509) | 1.215 (0.949-1.555) | 1.174 (0.910-1.515) |
| 26.5-27.5 | 478 | 1.253 (0.966-1.625) | 1.263 (0.964-1.655) | 1.160 (0.894-1.506) | 1.139 (0.870-1.491) |
| 27.5-28.5 | 361 | 1.378 (1.042-1.821) | 1.329 (0.993-1.778) | 1.657 (1.257-2.184) | 1.559 (1.173-2.073) |
| 28.5-29.5 | 256 | 1.217 (0.885-1.667) | 1.174 (0.841-1.632) | 1.674 (1.228-2.279) | 1.587 (1.151-2.184) |
| 29.5-30.5 | 197 | 1.512 (1.074-2.119) | 1.569 (1.097-2.234) | 1.844 (1.313-2.586) | 1.746 (1.230-2.474) |
| 30.5-31.5 | 116 | 1.287 (0.834-1.959) | 1.248 (0.794-1.936) | 1.949 (1.277-2.961) | 1.741 (1.124-2.685) |
| ≥31.5 | 270 | 1.193 (0.871-1.628) | 1.013 (0.729-1.403) | 3.124 (2.304-4.247) | 2.577 (1.881-3.536) |
| P for trend | | 0.089 | 0.241 | <0.001 | <0.001 |

Model 1: adjusted for age and sex.

Model 2: adjusted for age, sex, duration of DM, SBP, TG, LDL, HDL, HbA1c.

Table S8. Sensitivity analysis of WC and HC in relation to DR in men.

| | OR (95%CI) | | | |
|---|---|---|---|---|
| | Fifth of WC | | Fifth of HC | |
| | Model 4 | Model 5 | Model 4 | Model 5 |
| 1 (lowest) | | | | |
| 2 | 0.748 (0.564-0.992) | 0.982 (0.746-1.293) | 0.83 (0.635-1.084) | 0.845 (0.644-1.109) |
| 3 | 0.545 (0.395-0.75) | 0.847 (0.618-1.16) | 0.675 (0.51-0.893) | 0.697 (0.52-0.934) |
| 4 | 0.512 (0.36-0.729) | 0.942 (0.667-1.329) | 0.584 (0.429-0.794) | 0.609 (0.439-0.844) |
| 5 (highest) | 0.46 (0.296-0.712) | 1.133 (0.744-1.726) | 0.510 (0.358-0.724) | 0.546 (0.367-0.809) |
| P for trend | <0.001 | 0.559 | <0.001 | 0.001 |

Model 4: adjusted for age, sex, duration of DM, SBP, TG, LDL, HDL, HbA1c, BMI, WHR.

Model 5: adjusted for age, sex, duration of DM, SBP, TG, LDL, HDL, HbA1c, BMI, and mutually adjusted for WC and HC.
